# Evaluating Prominent Machine Translation Models for Kinyarwanda to English for Patient-Provider Conversations

**DOI:** 10.64898/2026.11.23.26368798

**Authors:** Hassan Mugabo, Riyaz Ur Rehman, Andy Liu, Matthew Manson, Justin Tuyisenge, Muhammed Semakula, Shera Potka, Richard Lester

## Abstract

**Background:** Linguistic barriers between patients and providers cause diagnostic error, poor adherence, and adverse events, and disproportionately affect speakers of low-resource languages. Kinyarwanda, spoken by more than 20 million people, has a small digital footprint, and the quality of off-the-shelf machine translation (MT) for its clinical dialogue is poorly documented.

**Methods:** We evaluated six MT systems (GPT-5, GPT-4o-mini, Gemini 2.5 Flash, Azure Translate, Google Translate, and NLLB-200) for Kinyarwanda-to-English patient-provider conversations. Using a professional reference corpus of 600 conversations, stratified across three pandemic periods and collected in Rwanda via the WelTel platform, we assessed automated metrics (semantic similarity, BLEU, chrF, and translation edit rate [TER]) and 0-100 quality ratings from three professional Kinyarwanda-English translators, with reliability quantified by intraclass correlation (ICC). De-identified secondary data were approved by UBC (H20-01031) and the Rwanda National Ethics Committee (100/RNEC/2020).

**Findings:** On the full 600-conversation cohort, GPT-4o-mini, GPT-5, and Gemini 2.5 Flash had closely clustered semantic similarity (0.877, 0.874, 0.873), well above Azure Translate (0.809) and NLLB-200 (0.754); Gemini 2.5 Flash led on BLEU (43.13) and the lowest TER (68.15). Translators ranked GPT-5 and Gemini 2.5 Flash highest (means 81.55 and 81.51), then GPT-4o-mini (72.67), Azure Translate (55.16), and NLLB-200 (39.05); average-rater reliability was moderate (ICC 0.594, rising to 0.701 on a balanced 85-conversation subset). Automated n-gram metrics understated the quality gap human raters perceived between large language models and conventional systems.

**Interpretation:** GPT-5 and Gemini 2.5 Flash are the strongest off-the-shelf systems for Kinyarwanda-English clinical translation and primary choices for human-in-the-loop use, with GPT-4o-mini a cost-efficient alternative. NLLB-200 underperformed, showing that models built for low-resource languages are not automatically suited to specialised clinical domains without fine-tuning. Human evaluation should be prioritised over automated metrics alone when comparing LLM-based translation.

## 1 Introduction

Communication is a cornerstone of medicine; the accuracy and nuance of information exchanged between a patient and a healthcare provider directly impacts diagnosis, treatment adherence, and overall patient outcomes [1]. Ensuring communication is effective across linguistically diverse settings is therefore fundamental to achieving equitable patient outcomes. Similarly, data collected for research interviews or survey inputs should include diversity of languages to ensure study populations are well represented. However, linguistic barriers often obstruct this vital exchange for millions of people worldwide, leading to bias in data collection, health disparities and adverse medical events [2].

To address these challenges, many organizations utilize professional translation services to translate clinical and research tools and data. In the United States, for example, the Department of Health and Human Services (HHS) and National Council on Interpreting in Health Care (NCIHC) developed national standards that encourage use of professional human translators [3]. However, researchers or healthcare providers serving populations that speak low-resource languages often face a dilemma: rely on the limited availability of human interpreters or turn to technology. Machine Translation (MT) systems have become increasingly accessible and powerful, offering a seemingly instantaneous solution. Yet, the quality of these systems can vary dramatically, and an inaccurate translation in a clinical setting can have severe consequences, ranging from misdiagnosis to incorrect medication administration[4].

With most Natural Language Processing (NLP) research today focusing on only about 20 of the world’s over 7,000 languages[5], a substantial technological gap remains. Kinyarwanda, a Bantu language spoken by over 20 million people primarily in Central African countries of Rwanda, Uganda, Democratic Republic of Congo and Burundi, is considered a low-resource language in the digital domain. This means it has a smaller data footprint for training natural language processing models compared to languages like English or Spanish, a challenge common to many African languages[6].

Despite the critical need, there is a lack of published research evaluating and comparing state-of-the-art MT models for languages with a small digital footprint, especially within the specialized domain of healthcare. In this context, accurate translation is essential for patient-provider communication, medical records, research questionnaires, interview transcripts, patient feedback, and beyond. This study aims to fill that gap by addressing the following research question: How do leading, publicly available machine translation models perform on the task of translating Kinyarwanda-English patient-provider conversations, and which model is most suitable for this context?

To answer this question, we conduct a quantitative evaluation of six models: Google Translate, Microsoft Azure Translate, Facebook/Meta’s No Language Left Behind (NLLB), Google’s Gemini 2.5 Flash, GPT5, and GPT-4o-mini. By comparing their output against a professionally translated reference set, we provide actionable insights for practitioners and a baseline for future research in this important area.

## 2 Methodology

Our evaluation framework is based on a parallel corpus of Kinyarwanda text message conversations between healthcare providers and patients professionally translated to English, a set of MT models accessed programmatically, and a suite of evaluation metrics calculated using standard libraries. The experiments were conducted within a Python 3 environment using Jupyter Notebooks, leveraging libraries such as Pandas for data manipulation and PyTorch for model inference.

### 2.1 Dataset

The basis for our evaluation is a parallel corpus comprising 600 conversations between patients and healthcare providers, which were collected in Rwanda during the COVID-19 pandemic via the bi-directional text messaging platform WelTel[7]. Each conversation is defined as any interactive text exchange that includes at least one patient message of three words or more and at least one message from a public health clinician or automated system. The corpus consists of source texts in Kinyarwanda and corresponding English reference translations. The translations were produced and then cross-checked by two professional human translators with expertise in both languages and the healthcare context. This professionally curated English text serves as the high-quality “gold standard” for all subsequent metric evaluations.

To capture the dynamic evolution of conversational topics and medical concerns over the course of the COVID-19 pandemic, the conversation database was divided into three chronological periods corresponding to the initial acute symptom-reporting phase, the peak-phase of healthcare strain, and the later vaccine rollout and preventive care period. A random sample without replacement of 200 conversations from each of the three temporal periods was then drawn, yielding a stratified main cohort of 600 conversations that is representative of the full range of pandemic-era exchanges.

Google Translate was evaluated on an initial pilot cohort of 400 conversations sampled chronologically prior to the adoption of the stratified design. Following the redesign of the sampling framework, re-running Google Translate on the full stratified cohort was not feasible due to resource constraints, and human evaluation had already been completed. Comparisons involving Google Translate are therefore restricted to the 85 conversations common to both cohorts.

### 2.2 Machine Translation Models and Implementation

We selected six widely used and technologically advanced MT systems. To ensure experimental consistency and minimize methodological bias, all models were applied to the same source corpus using a zero-shot approach (i.e., no task-specific fine-tuning or in-context examples). Where possible, batch translation was used to improve efficiency; individual request workflows were used where batch APIs were not available. Each model was accessed programmatically to translate the Kinyarwanda source text.

- NLLB-200 (distilled-600M): We utilized Meta AI’s “No Language Left Behind” model[8] via the Hugging Face transformers library[9]. Specifically, we loaded the facebook/ nllb-200-distilled-600M model and its corresponding tokenizer (AutoTokenizer). Translations were generated in batches of 8 on an NVIDIA RTX 4080 GPU, with the source language set to kin_Latn (Kinyarwanda) and the target language to eng_Latn (English).
- Gemini 2.5 Flash: We evaluated the translation capabilities of a general-purpose Large Language Model (LLM), specifically Google’s Gemini 2.5 Flash[10], by adapting it for the Kinyarwanda-to-English task via prompt engineering. This approach, conducted in a Python-based Google Colab environment using the google-genai SDK, relied on a carefully constructed zero-shot prompt to guide the model’s behavior. For each text segment, the model was given the following instructions:

> *You are a machine translation service. You will be given text in Kinyarwanda. Translate it to English. Do not add any extra commentary, notes, or apologies. Only provide the raw translated text. Kinyarwanda Text: “{text}” English Translation:*

While the Gemini API supports batch processing, our implementation performed the translation process sequentially for simplicity, processing one text segment at a time. The resulting English hypotheses were then saved to a new CSV file.

- Azure Translate: We used Microsoft’s Azure Translator service, a cloud-based Neural Machine Translation (NMT) system, to translate our Kinyarwanda test corpus into English. The process was automated with a Python script in a Google Colab environment, using the official azure-ai-translation-text SDK and the pandas library. After authenticating to the canadaeast service region with an API key, we implemented a batch processing workflow to manage request volume efficiently. Source texts from our dataset were sent in batches of 30 segments, with each API call explicitly defining Kinyarwanda (rw) as the source and English (en) as the target language. A crucial step was including a 90-second pause between each batch request to adhere to the service’s rate limits, ensuring the successful processing of the entire dataset. The resulting English translations were then systematically collected and saved to a new CSV file for subsequent metric evaluation.
- Google Translate: To establish another baseline, we employed the Google Cloud Translation API, a dedicated Neural Machine Translation (NMT) service. The implementation utilized the official google-cloud-translate Python library for the v3 version of the API, authenticated through a designated Google Cloud project. Our workflow processed the Kinyarwanda corpus sequentially, iterating through each text segment and sending an individual translation request to the API. Each request explicitly specified the source language as Kinyarwanda (rw) and the target language as English (en). Unlike other methods tested, no manual rate-limiting delays were implemented between requests; the process relied on the default API usage quotas. The resulting English translations were collected and saved to an output CSV file.
- GPT-4o-mini: OpenAI’s GPT-4o-mini was evaluated by accessing it programmatically via the OpenAI API using the openai Python SDK. The model was applied using a zero-shot translation prompt structurally identical to that used for Gemini, instructing the model to translate the provided Kinyarwanda text to English without additional commentary. Translations were performed sequentially, with each text segment submitted in an individual API request. The resulting English translations were collected and saved to a CSV file for metric evaluation.
- GPT-5: OpenAI’s GPT-5 was evaluated using the same OpenAI API workflow and zero-shot prompt as GPT-4o-mini, with the sole difference being the model identifier specified in the API request. This ensured that any performance differences between the two OpenAI models reflected model capability rather than methodological variation.

### 2.3 Evaluation Metrics and Implementation

We employed a combination of metrics to provide a holistic view of translation quality. Each metric is explained below with a non-technical description followed by its specific implementation details.

Metric Selection Rationale: We employed a combination of metrics to provide a holistic view of translation quality, focusing on semantic accuracy, fluency, and practical edit-distance. This multi-faceted approach ensures a robust evaluation. To maintain scientific validity, we excluded metrics like COMET, as its available pre-trained models do not officially support Kinyarwanda, which would produce unreliable scores. Our evaluation is therefore based on the following well established metrics.

- Semantic Similarity

*What it means:* This metric evaluates how well the machine translation preserves the core meaning of the human reference translation, even when different vocabulary or phrasing is used. It focuses on whether the two sentences convey the same essential message, which is paramount in a medical context where semantic errors can lead to dangerous clinical misinterpretations. The score ranges from 0 to 1, where 1 indicates an identical meaning and a score closer to 0 signifies that the meaning has been lost or distorted.

*Technical Implementation:* Our semantic similarity calculation was implemented in Python using the sentence-transformers library[11]. We first loaded the paraphrase-multilingual-mpnet-base-v2 model. [12]. The reference and hypothesis texts were then passed to the model.encode() function to convert them into tensor-based vector embeddings. Using these embeddings, we calculated the pairwise scores with the cos_sim function from the library’s util module. The final reported score is the mean of the diagonal of the resulting similarity matrix, which corresponds to the average similarity score for all translation pairs.

- BLEU (Bilingual Evaluation Understudy)

*What it means:* BLEU measures how much the machine’s translation overlaps with the professional human translation and works by counting matching individual words and short phrases[13]. A higher BLEU score suggests the translation is structurally similar to the reference and uses much of the same vocabulary. Following standard convention for reporting BLEU scores, the resulting score, which ranges from 0 to 1, is presented on a 0-100 scale for improved readability where a higher score is better.

*Technical Implementation:* The BLEU score was calculated using the sacrebleu library[14], which provides a standardized, corpus-level implementation.

- CHRF (Character n-gram F-score)

*What it means:* Popović introduced CHRF, a simple, language-independent automatic machine translation evaluation metric based on character n-gram F-scores, which shows a strong correlation with human judgments[15]. Similar to BLEU, CHRF also measures overlap, but it works at the character level instead of the word level. Following the same convention as BLEU, the resulting score, which ranges from 0 to 1, is presented on a 0-100 scale for improved readability, where a higher score is better.

*Technical Implementation:* The CHRF score was calculated using the sacrebleu library[14].

- TER (Translation Edit Rate)

*What it means:* It calculates the number of edits (additions, deletions, or substitutions) a human would need to make to fix the machine translation and make it perfect [16]. The scoring scale starts at 0 for a perfect translation and is unbounded, meaning scores can exceed 100. For this metric, a lower score is better, as it indicates the translation is closer to the human reference and requires less correction.

*Technical Implementation:* The TER score was calculated using the standardized implementation in the sacrebleu library[14].

The performance of each model across the suite of metrics was calculated by comparing their English outputs against the human reference translations.

### 2.4 Translation Quality Assessment by Humans

Three professional Kinyarwanda-English translators independently evaluated the translation quality of each model. Five models (GPT-5, Gemini 2.5 Flash, GPT-4o-mini, Azure Translate, and NLLB-200) were evaluated across the full 600-conversation main cohort. Google Translate was evaluated on the 85 overlapping conversations common to both the pilot and main cohorts, as described in Section 2.1. The translators were asked to assess the overall quality of each translation, with specific focus on the preservation of medical meaning, clarity and fluency, and consistency across conversation turns. They were also asked to flag translations that may be clinically misleading. A score between 0 and 100 was provided for each translation with reference to a standardized scoring scale (Table 1), which also served as the complete rating guidance provided to the translators. Inter-rater reliability was assessed using the intraclass correlation coefficient (ICC) for consistency, reported with 95% confidence intervals, with both single-rater and average-rater estimates computed.

**Table 1:** Scoring Guide Provided to Professional Kinyarwanda-English Translators.

| **Score Range** | **Interpretation** |
|---|---|
| 90–100: | Excellent translation; medically accurate, fluent, coherent, and safe |
| 70–89: | Mostly correct; minor issues, no impact on medical meaning |
| 50–69: | Noticeable errors; partial loss of meaning or ambiguity |
| 30–49: | Serious errors; misleading or inconsistent medical information |
| 0–29: | Unusable; meaning largely lost or potentially harmful |

### Statistical Analysis of Evaluator Reliability

To evaluate the reliability of the human expert panel, we calculated the Intraclass Correlation Coefficient (ICC) using a two-way mixed-effects consistency model, reporting both single-rater ICC(3,1) and average-rater ICC(3,k) metrics. Unlike standard correlation coefficients, the ICC consistency framework evaluates whether raters rank the machine translation models in the same relative order, successfully isolating true translation quality from individual rater harshness or leniency. Values were interpreted using standard benchmarks[17], where scores less than 0.5 indicate poor reliability, between 0.50 and 0.75 indicate moderate reliability, between 0.75 and 0.90 indicate good reliability, and scores greater than 0.90 indicate excellent reliability. All ICC metrics are reported with their corresponding 95% confidence intervals (CI) and p-values.

Probability values (p-values) are reported for all ICC metrics, with a significance threshold of p < 0.05. A p-value below this threshold indicates that the observed rater agreement is unlikely to have occurred by chance alone.

### 2.5 Assessment of Practical Considerations

Beyond quantitative metrics, we assessed each model’s practical usability for developers and healthcare implementers. This involved a qualitative evaluation of three key areas:

- Implementation Effort and Convenience: To systematically assess this, we defined and documented each model’s performance against three specific factors based on the direct experience of implementing them:

- **Setup and Dependencies:** We assessed the overhead required to begin using the model. This included whether it was a cloud-based API or required local setup, the need for specialized hardware (e.g., a GPU), and the necessary technical expertise (e.g., familiarity with libraries like Hugging Face).
- **API/Library Usability:** We evaluated the ease of processing our dataset. This included whether the API or library natively supported batch processing and how it handled rate-limiting (e.g., requiring manual pauses versus relying on default quotas).
- **Model Specialization:** We noted whether the model was a general-purpose tool that required specialized inputs or a dedicated translation-specific endpoint. A final, holistic rating (“High Effort,” “Moderate Convenience”, etc) was then assigned to each model based on the following factors:
- **Cost Structure**: We reviewed the publicly available pricing models for each service as of August 2025 to determine the financial costs associated with their use. Open-source models were evaluated based on their computational and hardware requirements rather than API fees.
- **Ecosystem and Language Support**: We examined the official documentation for each model to determine the total number of languages supported for translation, with a specific focus on counting the number of listed African languages to assess their suitability for diverse regional contexts.

### 2.6 Interpreting Evaluation Metrics

Before analyzing individual model performance, a framework for interpreting evaluation metrics is important. Different metrics measure distinct aspects of translation quality and can sometimes conflict (e.g., high semantic similarity but low lexical overlap with a reference). In the critical healthcare domain, we argue that not all metrics are equally important and propose a clear hierarchy.

Semantic Similarity is the primary metric. It addresses the key question: “Was the clinical meaning preserved?” A high score here is necessary for a successful translation, as semantic errors can lead to dangerous clinical misinterpretations.

Secondary to meaning is the quality and efficiency of the communication, best measured by TER (Translation Edit Rate). TER quantifies this effort by calculating the number of edits needed to perfect a translation. A semantically correct translation with a low TER is ideal, as it is both accurate and easy to understand, requiring minimal “correction effort” from the reader.

Tertiary metrics like BLEU and CHRF serve as indicators of fluency and lexical style. While valuable, a low score on these metrics is not necessarily a failure if the meaning is correct. It may simply indicate that the model used different, yet still valid, phrasing than our human reference. These metrics are most useful for assessing the naturalness of the output and for breaking ties between models with similar semantic performance.

## 3 Results

### 3.1 Quantitative Performance Metrics

Aggregated automated metric scores for the five models evaluated across the full 600-conversation main cohort are presented in **Table 2**. Google Translate is excluded from this primary comparison as it was evaluated on a separately sampled pilot cohort prior to the stratified design; its results are presented in the overlapping subset analysis in Section 3.3.

**Table 2:** Automated Translation Metrics — Full 600-Conversation Cohort (Google Translate excluded; see Section 3.3)

| Model | Semantic Similarity ↑ | BLEU ↑ | CHRF ↑ | TER ↓ |
|---|---|---|---|---|
| GPT-4o-mini | 0.8770 | 38.80 | 55.20 | 70.83 |
| GPT-5 | 0.8738 | 40.04 | 54.41 | 70.82 |
| Gemini 2.5 Flash | 0.8730 | 43.13 | 56.65 | 68.15 |
| Azure Translate | 0.8093 | 39.31 | 50.96 | 71.61 |
| NLLB-200 (600M) | 0.7543 | 14.09 | 26.53 | 129.11 |

Key: ↑ indicates that a higher score is better. ↓ indicates that a lower score is better.

As shown in Table 2, GPT-4o-mini recorded the highest Semantic Similarity score (0.8770), followed closely by GPT-5 (0.8738) and Gemini 2.5 Flash (0.8730), indicating strong meaning preservation across all three LLM-based systems. Gemini 2.5 Flash led on CHRF (56.65) and TER (68.15), suggesting its outputs require the least post-editing effort. Azure Translate achieved moderate performance across all metrics (Semantic Similarity: 0.8093; BLEU: 39.31), substantially outperforming NLLB-200 (600M), which scored lowest across all metrics.

### 3.2 Practical Considerations

In addition to performance on the translation task, the models differ significantly in their implementation effort, cost, and broader language support, which are critical factors for real-world deployment. These practical considerations are summarized in **Table 3**.

**Table 3:** Practical and Ecosystem Comparison of MT Models.

| Model | Implementation Effort & Convenience | Cost Model (as of Q3 2025) | African Language Support | Processing Time (600 dialogues) |
|---|---|---|---|---|
| Gemini 2.5 Flash | Moderate Convenience: Required prompt engineering to adapt the general-purpose model. While the API supports batching, our implementation processed texts sequentially. | Token-based with a free tier. Paid live API costs are per million tokens for input (\$0.50) and output (\$2.00). | Extensive support as part of its multimodal training. | ~55 minutes(extrapolated from 85 translations) |
| Azure Translate | High Convenience: Natively supports batch processing. A manual pause between batches was needed. | Generous free tier of 2 million characters/month. Pay-as you-go at \$10 per million characters. | Very strong; about 27 African languages supported. | ~33 minutes |
| Google Translate | Highest Convenience: Relied on default API usage quotas without needing manual rate-limiting. | Free tier of 500,000 characters/month. Payas-you-go at \$20 per million characters. | Industry-leading; about 59 African languages supported. | ~5 minutes |
| NLLB-200 | High Effort: Requires local setup, GPU hardware, and technical expertise with the Hugging Face library. | Free & Open-Source (Cost is in hardware/computation). | Excellent; over 50 African languages supported. | ~17 minutes (on RTX 4080 GPU) |
| GPT-4o-mini | Moderate Convenience: Accessed via the OpenAI API using a zero-shot prompt in a sequential per-request workflow. No specialized hardware required; straightforward to implement. | Token-based; no free tier for API access. Input priced at \$0.15 per million tokens, output at \$0.60 per million tokens. Estimated cost for this corpus was less than 0.10 usd. | Broad multilingual support as a general-purpose LLM; number of formally supported African languages is not explicitly documented. | ~110 minutes |
| GPT-5 | Moderate Convenience: Accessed via the OpenAI API using the identical zero-shot workflow as GPT-4o-mini, with only the model identifier changed. Implementation overhead is equivalent. | Token-based; no free tier for API access. Input priced at \$2.00 per million tokens, output at \$8.00 per million tokens. Estimated cost for this corpus was approximately \$1.00. | Broad multilingual support as a general-purpose LLM; formal African language coverage not explicitly catalogued. | ~90 minutes |

While NLLB-200 offers a cost-free, open-source solution with extensive African language support, it demands the most significant technical expertise and computational resources to deploy.

Among the commercial APIs, the cost structures vary significantly. For a one-time translation of our corpus ( 0.5 million characters), the direct costs were negligible. Azure Translate, with the most generous free tier at 2 million characters per month, processed the entire corpus for $0.00. Google Translate’s free tier of 500,000 characters per month also meant the cost for our run was $0.00. Gemini’s token-based pricing is distinct, but the estimated cost for our specific run was also minimal at approximately $0.25.

In terms of implementation and speed, Google Translate was the most straightforward and fastest, processing the entire corpus in about 5 minutes by leveraging its default API quotas. Azure was also highly convenient, though it required a manual pause between batches and took approximately 33 minutes. Gemini, despite its top performance, required the most hands-on effort of the commercial APIs due to prompt engineering and a sequential implementation, resulting in a processing time of about 55 minutes. The local NLLB-200 model was faster than the sequential Gemini and batched Azure implementations, completing the task in roughly 17 minutes on a GPU

However, from a post-editing perspective, Gemini 2.5 Flash recorded the lowest TER (68.15), indicating it would require the least post-editing effort, while GPT-5 (70.82), GPT-4o-mini (70.83), and Azure Translate (71.61) were closely clustered, suggesting broadly comparable correction overhead among the three

### 3.3 Human Evaluation and Inter-Rater Reliability

To validate the automated metrics, three professional translators independently evaluated the translation quality of each model. For the full 600-conversation dataset, single-rater ICC was 0.327 (95% CI [0.30, 0.35], p < 0.001) and average-rater ICC was 0.594 (95% CI [0.57, 0.62], p < 0.001). The average-rater estimate, which reflects the reliability of the consensus panel score used in subsequent analyses, indicates moderate agreement per standard benchmarks[17]

Table 4 presents statistics by individual rater. Rater 2 applied systematically harsher scores, while Rater 3 applied more generous scores, yet both maintained consistent relative model rankings. Table 5 presents the consolidated human evaluation scores (mean of all three raters) for the full dataset.

**Table 4:** Individual Rater Statistics — Full 600-Conversation Dataset.

| Rater | Model | Mean | Median | Std Dev |
|---|---|---|---|---|
| Rater 1 | GPT-5 | 75.08 | 75.0 | 10.31 |
|  | Gemini 2.5 Flash | 79.04 | 80.0 | 10.09 |
|  | GPT-4o-mini | 77.48 | 78.0 | 8.36 |
|  | Azure Translate | 73.78 | 75.0 | 7.87 |
|  | NLLB-200 | 58.78 | 65.0 | 15.82 |
| Rater 2 | GPT-5 | 72.54 | 70.0 | 20.35 |
|  | Gemini 2.5 Flash | 70.52 | 70.0 | 21.97 |
|  | GPT-4o-mini | 49.88 | 50.0 | 17.25 |
|  | Azure Translate | 43.41 | 50.0 | 30.92 |
|  | NLLB-200 | 35.93 | 30.0 | 27.74 |
| Rater 3 | GPT-5 | 97.02 | 100.0 | 10.55 |
|  | Gemini 2.5 Flash | 95.34 | 100.0 | 15.20 |
|  | GPT-4o-mini | 90.69 | 100.0 | 18.88 |
|  | Azure Translate | 48.18 | 70.0 | 25.93 |
|  | NLLB-200 | 8.08 | 0.0 | 20.65 |

**Table 5:** Consolidated Human Evaluation Scores − Full 600-Conversation Dataset.

| Model | Mean Score | Median Score | Std Dev |
|---|---|---|---|
| GPT-5 | 81.55 | 85.0 | 18.18 |
| Gemini 2.5 Flash | 81.51 | 85.0 | 19.40 |
| GPT-4o-mini | 72.67 | 75.0 | 23.02 |
| Azure Translate | 55.16 | 70.0 | 27.20 |
| NLLB-200 | 39.05 | 50.0 | 29.19 |

### 3.4 Overlapping Subset Analysis (85 Conversations)

To enable a completely balanced comparison including Google Translate, a secondary evaluation was conducted restricted to the 85 conversations present in both the pilot cohort and the stratified main cohort. On this subset, inter-rater agreement was higher than on the full dataset. This higher agreement on the smaller subset likely reflects the reduced conversational diversity of the 85 overlapping conversations compared to the full stratified cohort, which may make relative model quality easier for raters to distinguish consistently. Single-rater ICC was 0.439 (95% CI [0.39, 0.49], p < 0.001) and average-rater ICC was 0.701 (95% CI [0.65, 0.74], p < 0.001). The average-rater ICC indicates good reliability for the consensus panel score.

Table 6 presents the consolidated human evaluation scores on this balanced subset, and Table 7 presents the corresponding automated metric scores. The model rankings on human scores remained broadly consistent with the full dataset: GPT-5 and Gemini 2.5 Flash led the ranking (84.95 and 83.34, respectively), while NLLB-200 scored lowest (29.25). Two notable shifts were observed. First, Azure Translate’s human score increased substantially from 55.16 (full cohort) to 71.45 (balanced subset), indicating that the 85-conversation overlap did not capture the specific dialogue types where Azure Translate underperforms on the wider corpus. Second, NLLB-200’s score declined further from 39.05 to 29.25, confirming its difficulty with the vocabulary and conversational structure present in these exchanges.

**Table 6:** Consolidated Human Evaluation Scores — 85-Conversation overlapping Subset.

| Model | Mean Score | Median Score | Std Dev |
|---|---|---|---|
| GPT-5 | 84.95 | 88.0 | 14.26 |
| Gemini 2.5 Flash | 83.34 | 85.0 | 15.90 |
| GPT-4o-mini | 72.43 | 76.5 | 25.16 |
| Google Translate | 72.07 | 83.0 | 24.80 |
| Azure Translate | 71.45 | 70.0 | 15.56 |
| NLLB-200 | 29.25 | 30.0 | 29.11 |

**Table 7:** Automated Translation Metrics — 85-Conversation overlapping Subset.

| Model | Semantic Similarity ↑ | BLEU ↑ | CHRF ↑ | TER ↓ |
|---|---|---|---|---|
| Gemini 2.5 Flash | 0.8385 | 36.76 | 56.82 | 89.58 |
| Google Translate | 0.8526 | 36.65 | 58.16 | 89.87 |
| GPT-4o-mini | 0.8528 | 35.54 | 57.35 | 85.13 |
| GPT-5 | 0.8283 | 34.87 | 55.12 | 90.34 |
| Azure Translate | 0.7495 | 30.55 | 54.80 | 92.51 |
| NLLB-200 | 0.7113 | 5.24 | 20.20 | 198.01 |

A comparison of the automated and human rankings on this subset show a divergence. On automated metrics, GPT-4o-mini and Google Translate led on semantic similarity (0.8528 and 0.8526 respectively), while GPT-5 recorded the lowest semantic similarity among the top models (0.8283). Yet human evaluators ranked GPT-5 highest (84.95) and Gemini 2.5 Flash second (83.34), both substantially above Google Translate (72.07) despite its stronger automated scores. This pattern is further illustrated by BLEU: automated metrics rated Gemini 2.5 Flash (36.76), Google Translate (36.65), and GPT-4o-mini (35.54) as nearly equivalent, yet human evaluators rated Gemini 11.27 points higher than Google Translate (83.34 vs. 72.07). This reversal across both embedding and n-gram metrics suggests that advanced LLMs generate more natural, conversationally fluent translations that diverge lexically from the reference text, causing automated metrics to systematically underestimate their quality relative to human judgment. Researchers evaluating LLM-based translation should therefore prioritise human evaluation over automated metrics alone when comparing LLMs against traditional NMT systems.

## 4 Discussion

### 4.1 GPT-5 and Gemini 2.5 Flash Lead the Metrics

Across both the full 600-conversation cohort and the 85-conversation overlapping subset, GPT-5 and Gemini 2.5 Flash emerged as the consistently top-performing models, demonstrating a level of robustness not observed in the other systems evaluated. On the full cohort, both models achieved near-identical consolidated human evaluation scores (GPT-5: 81.55; Gemini 2.5 Flash: 81.51), and this ranking was maintained on the overlapping subset (GPT-5: 84.95; Gemini 2.5 Flash: 83.34). Gemini 2.5 Flash additionally recorded the lowest TER score on the full cohort (68.15), indicating that its translations required the least post-editing effort relative to the reference. These results show that GPT-5 and Gemini 2.5 Flash are the primary recommendations for high-stakes clinical translation tasks where accuracy and fluency are important.

### 4.2 N-gram Metrics alone failed to accurately capture LLM Translation Quality

We find that traditional automated metrics, specifically BLEU, CHRF, and TER, are poorly suited to evaluating the output of advanced LLMs when compared against translations produced by conventional neural MT systems. On the 85-conversation overlapping subset, automated metrics rated Gemini 2.5 Flash (BLEU: 36.76), Google Translate (BLEU: 36.65), and GPT-4o-mini (BLEU: 35.54) as nearly equivalent. Human evaluators, however, rated Gemini 2.5 Flash 11.27 points higher than Google Translate (83.34 vs. 72.07), and GPT-5 12.88 points higher (84.95 vs. 72.07), despite Google Translate achieving a higher BLEU score than GPT-5 (36.65 vs. 34.87), and despite GPT-5 recording the lowest semantic similarity among the top models on this subset (0.8283), below Google Translate (0.8526) and GPT-4o-mini (0.8528). LLMs generate translations that are more natural and conversationally fluent, paraphrasing instead of direct word-for-word output. Because BLEU and CHRF measure lexical overlap with a single reference translation, they penalise this variance even when the meaning is superior.

### 4.3 Azure Translate showed moderate but Inconsistent performance

Azure Translate occupies a middle tier in this evaluation, but its cross-subset variability warrants careful consideration. On the full 600-conversation cohort, Azure Translate received a consolidated human evaluation score of 55.16, placing it substantially below GPT-4o-mini (72.67). On the 85-conversation overlapping subset, however, its score increased to 71.45, placing it statistically on par with both GPT-4o-mini (72.43) and Google Translate (72.07). This 16-point swing suggests that Azure Translate’s performance is sensitive to the specific types of dialogue present in the evaluation set, and that certain conversational structures or vocabulary present in the wider corpus expose translation weaknesses not evident in the overlap subset. Practitioners should therefore exercise caution when interpreting Azure Translate evaluations conducted on small or non-representative samples: while it may appear competitive on a narrow subset, its performance can degrade significantly across a broader and more diverse corpus. For applications requiring consistent quality across varied patient-provider exchanges, a more robust system such as GPT-5 or Gemini 2.5 Flash is preferable..

### 4.4 Google Translate showed speed and convenience at a Measured Quality Cost

Google Translate was evaluated exclusively on the 85 overlapping conversations common to both cohorts, achieving a human evaluation score of 72.07, placing it in the middle tier alongside GPT-4o-mini and Azure Translate on this subset. Its automated metrics were competitive: semantic similarity (0.8526), BLEU (36.65), and CHRF (58.16) were all broadly comparable to the top LLMs on the same subset, though this comparability reflects the limitation of n-gram metrics discussed in Section 4.2 rather than equivalent fluency or naturalness. Google Translate’s primary advantage remains its speed and ease of access: it processed our corpus in approximately 5 minutes with no manual rate-limiting. Its widespread availability and easily accessible web application make it a convenient tool for quick translations, particularly when dealing with a smaller number of text segments. However, it should be noted that human evaluators placed it approximately 12 points below GPT-5 and 11 points below Gemini 2.5 Flash on the same conversations, and that its outputs may require more manual review in high-stakes clinical documentation contexts.

### 4.5 NLLB-200 Underperformed

The NLLB-200 distilled model (600M parameters) performed poorly across all evaluation dimensions. On the full 600-conversation cohort, it recorded the lowest semantic similarity (0.7543), the lowest BLEU score (14.09), and the highest TER (129.11), indicating that its translations were frequently inaccurate and would require substantial post-editing to be usable. Human evaluators awarded it a mean score of 39.05 on the full cohort and 29.25 on the overlapping subset. Its performance on the balanced subset was notably worse than on the full cohort, further confirming that this model struggles with the specific vocabulary and conversational register present in Kinyarwanda patient-provider exchanges. This underperformance is likely attributable to a combination of the model’s smaller parameter count (the distilled 600M variant was used rather than the full 1.3B or 3.3B models) and the absence of in-domain medical training data. As an open-source system, NLLB-200 does retain the potential for domain adaptation, and fine-tuning on a dedicated Kinyarwanda medical corpus remains a promising avenue for future work.

## 4 Limitations and Future Work

First, our implementation choices, such as using a sequential process for Gemini, influenced the processing times; future work could explore the speed implications of fully optimized batch processing for all APIs.

Second, future work could also involve a blind review of the translations by medical professionals and Kinyarwanda speakers to assess their practical usability and safety. Furthermore, investigating the potential of fine-tuning models like NLLB on a dedicated Kinyarwanda medical corpus could be a promising avenue for developing a truly specialized and high-quality translation tool.

Third, Google Translate was evaluated on an earlier pilot cohort and could not be re-run on the full stratified sample due to resource constraints, limiting its direct comparability with the other five models.

## 5 Conclusion

In response to our research question, this study establishes a performance hierarchy among six off-the-shelf MT systems for Kinyarwanda-English healthcare translation, combining automated metrics with independent human expert evaluation to provide a multi-dimensional assessment.

GPT-5 and Gemini 2.5 Flash are the top-performing systems across both automated and human evaluation criteria, achieving the highest human quality scores on the full cohort (81.55 and 81.51, respectively) and maintaining those rankings on the overlapping subset. For practitioners where translation quality is the highest priority, either system represents a well-supported primary choice. Gemini 2.5 Flash additionally offers the lowest post-editing effort (TER: 68.15) among the full-cohort models, making it particularly attractive for workflows involving human review.

GPT-4o-mini achieved the highest automated semantic similarity score on the full cohort (0.8770) and a human evaluation score of 72.67, placing it as a capable and cost-efficient alternative where budget constraints preclude the use of higher-tier models.

Azure Translate and Google Translate occupy a middle tier on human evaluation, with competitive automated metrics but lower fluency ratings from professional translators. Azure Translate’s performance variability across evaluation subsets (55.16 on the full cohort vs. 71.45 on the overlapping subset) suggests it should be validated on a representative sample before broad deployment. Google Translate remains the fastest and most accessible option for near-instantaneous use cases.

NLLB-200 underperformed across all metrics, underscoring that models designed for low-resource languages are not automatically suited to specialised clinical domains without fine-tuning.

Ultimately, this evaluation serves a data-driven guide for healthcare providers, NGOs, global health, health equity researchers, technology implementers etc. By choosing the right tool and ensuring validation, they can dramatically speed up and reduce the cost of translation, effectively bridge the communication gap and ensuring that Kinyarwanda-speaking patients receive safer and higher-quality care.

## Data Availability

The data underlying this study cannot be shared publicly because they consist of patient-provider health conversations collected in Rwanda that contain sensitive personal health information. Public release is restricted by the terms of the original data collection protocol approved by the Rwanda National Ethics Committee (100/RNEC/2020) and the University of British Columbia Research Ethics Board (H20-01031). Data are available from the Rwanda Biomedical Centre, Research, Innovation and Data Science Division (contact via), for researchers who meet the criteria for access to confidential data, subject to approval and a data-use agreement.

## Declarations

### Ethics approval

University of British Columbia Research Ethics Board (certificate H20-01031) and Rwanda National Ethics Committee (100/RNEC/2020). De-identified secondary data; all participants provided informed consent at the original data collection, and secondary use of de-identified data was covered under the existing protocols. No patients were directly involved in the design of this benchmark study.

### Contributors

H.M.: Data curation, Resources, Validation, and Writing - review and editing. R.U.R.: Conceptualization, Methodology, Software, Formal analysis, and Writing - original draft. A.L.: Software and Validation. M.M.: Investigation, Data curation, and Formal analysis. J.T.: Data curation and Validation. M.S.: Methodology and Formal analysis. S.P.: Formal analysis, Visualization, and Writing - review and editing. R.T.L.: Conceptualization, Supervision, Funding acquisition, Resources, and Writing - review and editing. All authors had access to the study data, contributed to the interpretation of results, and read and approved the final manuscript.

### Data sharing

The de-identified machine-translation outputs and the analysis scripts used to generate the results in this Article are available from the corresponding author on reasonable request. The underlying WelTel COVID-19 patient-provider dialogue corpus contains sensitive health data and is subject to the data-sharing terms of the original collection protocol; access requires approval from the Rwanda Biomedical Centre and a data-use agreement, and cannot be made openly available. No other supporting documents are available.

